# Real-life validation of the Panbio COVID-19 Antigen Rapid Test (Abbott) in community-dwelling subjects with symptoms of potential SARS-CoV-2 infection

**DOI:** 10.1101/2020.10.16.20214189

**Authors:** Hendrik Gremmels, Beatrice M.F. Winkel, Rob Schuurman, Andert Rosingh, Nicolette A.M. Rigter, Olga Rodriguez, Johan Ubijaan, Annemarie M.J. Wensing, Marc J.M. Bonten, L.Marije Hofstra

**Author notes:** these authors contributed equally (Hendrik Gremmels, Beatrice M.F. Winkel). Corresponding author: L.M. Hofstra.

## Abstract

**Background:** RT-qPCR is the reference test for identification of active SARS-CoV-2 infection, but is associated with diagnostic delay. Antigen detection assays can generate results within 20 minutes and outside of laboratory settings. Yet, their diagnostic test performance in real life settings has not been determined.

**Methods:** The diagnostic value of the Panbio™ COVID-19 Ag Rapid Test (Abbott), was determined in comparison to RT-qPCR (Seegene Allplex) in community-dwelling mildly symptomatic subjects in a medium (Utrecht, the Netherlands) and high endemic area (Aruba), using two concurrently obtained nasopharyngeal swabs.

**Findings:** 1367 and 208 subjects were enrolled in Utrecht and Aruba, respectively. SARS-CoV-2 prevalence, based on RT-qPCR, was 10.2% (n=139) and 30.3% (n=63) in Utrecht and Aruba respectively. Specificity of the Panbio™ COVID-19 Ag Rapid Test was 100% (95%CI: 99.7-100%) in both settings. Test sensitivity was 72.6% (95%CI: 64.5-79.9%) in the Netherlands and 81.0% (95% CI: 69.0–89.8%) in Aruba. Probability of false negative results was associated with RT-qPCR Ct-values, but not with duration of symptoms. Restricting RT-qPCR test positivity to Ct-values <32 yielded test sensitivities of 95.2% (95%CI: 89.3-98.5%) in the Netherlands and 98.0% (95%CI: 89.2–99.95%) in Aruba.

**Interpretation:** In community-dwelling subjects with mild respiratory symptoms the Panbio™ COVID-19 Ag Rapid Test had 100% specificity, and a sensitivity above 95% for nasopharyngeal samples when using Ct values <32 cycles as cut-off for RT-qPCR test positivity. Considering short turnaround times, user friendliness, low costs and opportunities for decentralized testing, this test can improve our efforts to control transmission of SARS-CoV-2.

**Funding:** UMCU and LABHOH, Aruba

## Introduction

The SARS-CoV-2 pandemic has extensive impact on healthcare globally, with over 37 million confirmed cases and currently more than one million deaths.^1^ Rapid diagnosis of SARS-CoV-2 infection and subsequent contact tracing are essential in the containment of transmission.^2^ The reference test for detection of acute SARS-CoV-2 infection is reverse transcriptase quantitative polymerase chain reaction (RT-qPCR).^3^ RT-qPCR requires the use of expensive laboratory instrumentation as well as dedicated lab supplies and trained personnel, which causes significant challenges to generate sufficient testing capacity and short turnaround times.

Lateral Flow Assay (LFA)-based point of care tests (POCT) for rapid antigen detection using antibodies are cheap, simple to perform, do not require laboratory instrumentation and generate results within 20 minutes. Several different rapid antigen tests have been developed with usually high specificity but varying sensitivity^4–9^. These tests have the potential to alter testing strategies worldwide.^7^ Recently the WHO approved the first rapid diagnostic POCT and initiated a global partnership to pledge 120 million tests to low- and middle-income countries.^10^ However, the diagnostic performance of individual POCTs in real-life community settings is unknown.^9^

We evaluated the Abbott Panbio™ COVID-19 Ag rapid test in community testing locations in both a medium- and high endemic population and compared results to RT-qPCR and determined associations with duration of symptoms and risk of exposure.

## Materials and methods

### Populations and Study Period

All individuals visiting COVID-19 community testing centers, located at the University Medical Center Utrecht (UMCU) in the Netherlands (September 22^nd^ to October 6^th^) and the Horacio Oduber Hospital on Aruba (September 23^rd^ to October 9^th^), aged 16 and over were asked to participate in the evaluation. In both study sites, subjects were first sampled for routine RT-qPCR testing, using a combined throat/nasopharyngeal swab. Study participants received an additional nasopharyngeal swab. Participants at the UMCU study site were asked to fill out a questionnaire regarding (onset of) symptoms and risk of exposure to SARS-CoV-2.

### Diagnostic tests

#### RT-qPCR

After collection, swabs were transferred into 3 ml Universal transport medium until further processing. Nucleic acid extraction, RT-PCR and results interpretation were performed according the instructions of the manufacturer (Seegene, South-Korea). In short, RNA was isolated and purified using the MagC extraction kit (Seegene, South-Korea) on an automatic nucleic acid extractor Hamilton MicroLAB StartLET (Bonaduz, Switzerland). Subsequently, cDNA was generated and amplification was performed in a single tube assay using the Allplex 19-nCoV multiplex platform for detection of SARS-CoV-2 (Seegene, South-Korea), and results were interpreted with Seegene Viewer data analysis software. The assay uses fluorescent Taqman® probes for three SARS-CoV-2 genes (E-, N-, and RdRP-gene). Amplification and detection were performed for 45 cycles on a Biorad CFX96 thermocycler (Biorad Laboratories, the Netherlands). A positive result was defined as amplification of any of the three SARS-CoV-2 genes.

#### LFA

The Panbio™COVID-19 Ag rapid test device by Abbott (Lake Country, IL, U.S.A) is a membrane-based immunochromatography assay which detects the nucleocapsid protein of SARS-CoV-2 in nasopharyngeal samples. Collected swabs were transferred into dedicated sample collection tubes containing a sampling buffer. Collected samples were subsequently processed in a level 2 biosafety cabinet in accordance with the manufacturer’s protocol, within 2 hours of sample collection. Test results were recorded after 15 minutes of assay initiation, by two independent observers (blinded to each other and to the PCR results). Single lot LFA testing devices were used: lot 41ADF011A.

### Ethical Approval

The medical research ethics committee (MREC) of Utrecht decided the study is not subject to the Medical Research Involving Human Subjects Act (WMO) and did not require full review by an accredited MREC. The ethical committee of the hospital board of Aruba approved the study. All participants have provided written informed consent.

### Statistical analysis

Population characteristics are reported as mean (SD) or median [IQR] values. Specificity and sensitivity with 95% confidence intervals, and positive and negative predictive value of the LFA were calculated using the RT-qPCR results as reference test. Factors associated with LFA results were determined using logistic regression, using Nagelkerke’s pseudo R^2^ as a measure of goodness-of-fit. Data was analysed using the free, open-source software environment R ^11^.

## Results

### Population Characteristics

At the UMCU study site 1369 subjects were included, of which 139 tested positive for SARS-CoV-2 by RT-qPCR (prevalence: 10.2%). The mean (SD) Ct values for E-gene, N-gene and RdRP-gene were 24.74 (5.73), 27.51 (6.01) and 26.35 (5.60), respectively.

At the Aruba study site 208 subjects were included, of which 63 tested positive for SARS-CoV-2 (prevalence: 30.3%). The mean (SD) Ct values for E-gene, N-gene and RdRP-gene at the Aruba site were 25.69 (5.96), 26.56 (6.41) and 26.26 (6.36), respectively.

Estimated participation rates were 50% at the UMCU study site and 25% at the Aruba study site. Samples from two participants were excluded, due to inappropriate application of the nasopharyngeal swab and laboratory mislabelling. Data on symptoms were missing from nine subjects and data on duration of symptoms from 201 subjects (14.7% of total subjects, 11.9% of SARS-CoV-2 positive subjects, p=0.439).

Individuals at the UMCU study site were more often female (61.7%) and were largely between 20 to 50 years of age (Table 1). Nearly all individuals reported symptoms (97.3%), most frequently coryza (69.0%), sore throat (66.3%) and cough (57.1%). Duration of symptoms were 1-3 days in 387 subjects (33.2%), 4-7 days in 560 subjects (48.0%), and more than a week in 191 subjects (16.4%). Of all individuals, 17% reported prior contact with a confirmed SARS-CoV-2 positive individual.

**Table 1:** Population characteristics (UMCU study site).

|  | *Total* | *SARS-CoV-2*<br>*Negative\** | *SARS-CoV-2*<br>*Positive\** | *p-Value* |
| --- | --- | --- | --- | --- |
| *n* | 1367 | 1228 | 139 |  |
| *Sex = F (%)* | 844 (61.7) | 772 (63.1) | 72 (52.2) | **0.016** |
| *Age (median [IQR])* | 36.41 [27.0, 49.6] | 36.59 [27.34, 49.60] | 33.73 [23.42, 49.29] | **0.034** |
| *Contact with Confirmed Positive* | 233 (17.0) | 185 (15.1) | 48 (34.5) | **<0.001** |
| *Asymptomatic* | 37 (2.7) | 34 (2.8) | 3 (2.2) | 0.875 |
| *Duration of symptoms:* |  |  |  | **0.019** |
| *1-3 days* | 387 (33.2) | 353 (33.6) | 34 (29.6) |  |
| *4-7 days* | 560 (48.0) | 493 (46.9) | 67 (58.3) |  |
| *>7 days* | 191 (16.4) | 181 (17.2) | 10 (8.7) |  |
| *Symptoms:* |  |  |  |  |
| *Fever (%)* | 221 (16.2) | 170 (13.9) | 51 (36.7) | **<0.001** |
| *Chills (%)* | 279 (20.4) | 225 (18.5) | 54 (38.8) | **<0.001** |
| *Sore Throat (%)* | 907 (66.3) | 839 (68.8) | 68 (48.9) | **<0.001** |
| *Cough (%)* | 780 (57.1) | 692 (56.8) | 88 (63.3) | 0.165 |
| *Shortness of Breath (%)* | 274 (20.0) | 250 (20.5) | 24 (17.3) | 0.429 |
| *Coryza (%)* | 943 (69.0) | 867 (71.1) | 76 (54.7) | **<0.001** |
| *Altered Smell or Taste (%)* | 202 (14.8) | 159 (13.0) | 43 (30.9) | **<0.001** |
| *General Malaise (%)* | 365 (26.7) | 317 (26.0) | 48 (34.5) | **0.041** |
| *Abdominal Pain (%)* | 108 (7.9) | 97 (8.0) | 11 (7.9) | 1.000 |
| *Vomiting (%)* | 32 (2.3) | 26 (2.1) | 6 (4.3) | 0.189 |
| *Nausea (%)* | 115 (8.4) | 100 (8.2) | 15 (10.8) | 0.380 |
| *Diarrhoea (%)* | 137 (10.0) | 122 (10.0) | 15 (10.8) | 0.887 |
| *Headache (%)* | 601 (44.0) | 520 (42.7) | 81 (58.3) | **0.001** |
| *Rash (%)* | 25 (1.8) | 22 (1.8) | 3 (2.2) | 1.000 |
| *Eye Infection (%)* | 31 (2.3) | 26 (2.1) | 5 (3.6) | 0.426 |
| *Muscle Ache (%)* | 247 (18.1) | 195 (16.0) | 52 (37.4) | **<0.001** |
| *Joint Ache (%)* | 111 (8.1) | 83 (6.8) | 28 (20.1) | **<0.001** |
| *Tiredness (%)* | 565 (41.3) | 491 (40.3) | 74 (53.2) | **0.004** |
| *Reduced Appetite (%)* | 132 (9.7) | 111 (9.1) | 21 (15.1) | **0.035** |
| *E-gene Ct-value (mean (SD))* |  |  | 24.74 (5.73) |  |
| *N-gene Ct-value (mean (SD))* |  |  | 27.51 (6.01) |  |
| *RdRP-gene Ct-value (mean (SD))* |  |  | 26.35 (5.60) |  |

\* based on RT-qPCR results

Compared to subjects who tested negative for SARS-CoV-2 in RT-qPCR, SARS-CoV-2 positive subjects were younger, more likely male, more frequently had prior contact with aconfirmed SARS-CoV-2 positive individual, and more frequently reported fever, chills, an altered sense of smell or taste, or joint- or muscle ache. The most frequently reported symptoms, coryza and sore throat, were negatively associated with detection of SARS-CoV-2 by RT-qPCR.

### LFA Results

At the UMCU study site, 101 subjects tested positive by LFA yielding an overall sensitivity of 72.6% (95% CI: 64.5 – 79.9%) (Table 2). False positive LFA results were not observed (specificity 100%, 95% CI: 99.7 – 100%). Similar results were obtained at the Aruba study site, with an overall sensitivity of 81.0% (95% CI: 69.0 – 89.8%) and specificity of 100% (95% CI:97.5 – 100%) (Table 2). We observed no interrater variability in interpretation of test bands and no bands were classified as unclear by the independent observers.

**Table 2:** Test characteristics of the LFA compared to the RT-qPCR for the UMCU study site and the Aruba study site.

| Study site | LFA result | PCR result |  |  | Specificity | Sensitivity |
| --- | --- | --- | --- | --- | --- | --- |
|  |  | Positive (Ct<32) | Positive (Ct ≥32) | Negative |  |  |
| UMCU | Positive | 101 | 0 | 0 | 100% | Overall: 72.6% (64.5-79.9) |
|  | Negative | 5 | 33 | 1228 | (99.7-100) | Ct<32: 95.2% (89.3-98.5) |
| Aruba | Positive | 48 | 3 | 0 | 100% | Overall: 81.0% (69.0-89.9) |
|  | Negative | 1 | 11 | 145 | (97.5-100) | Ct<32: 98.0% (89.2-99.95) |

Sensitivity and specificity are reported with 95% CI.

False negative LFA results were mostly observed in subjects with high RT-qPCR Ct values,reflecting low viral load levels in nasopharyngeal material (Figure 1). Using logistic regression, the likelihood of a false negative LFA result was associated with the RT-qPCR Ct value (R^2^ = 0.77, p<0.0001, Figure 2). When defining RT-qPCR Ct positivity on a cut-off Ct value of 32 LFA sensitivity was 95.2% (95% CI: 89.3 – 98.5%) at the UMCU study and 98.0% (89.2 – 99.95%) at the Aruba study site.

**Figure 1.**
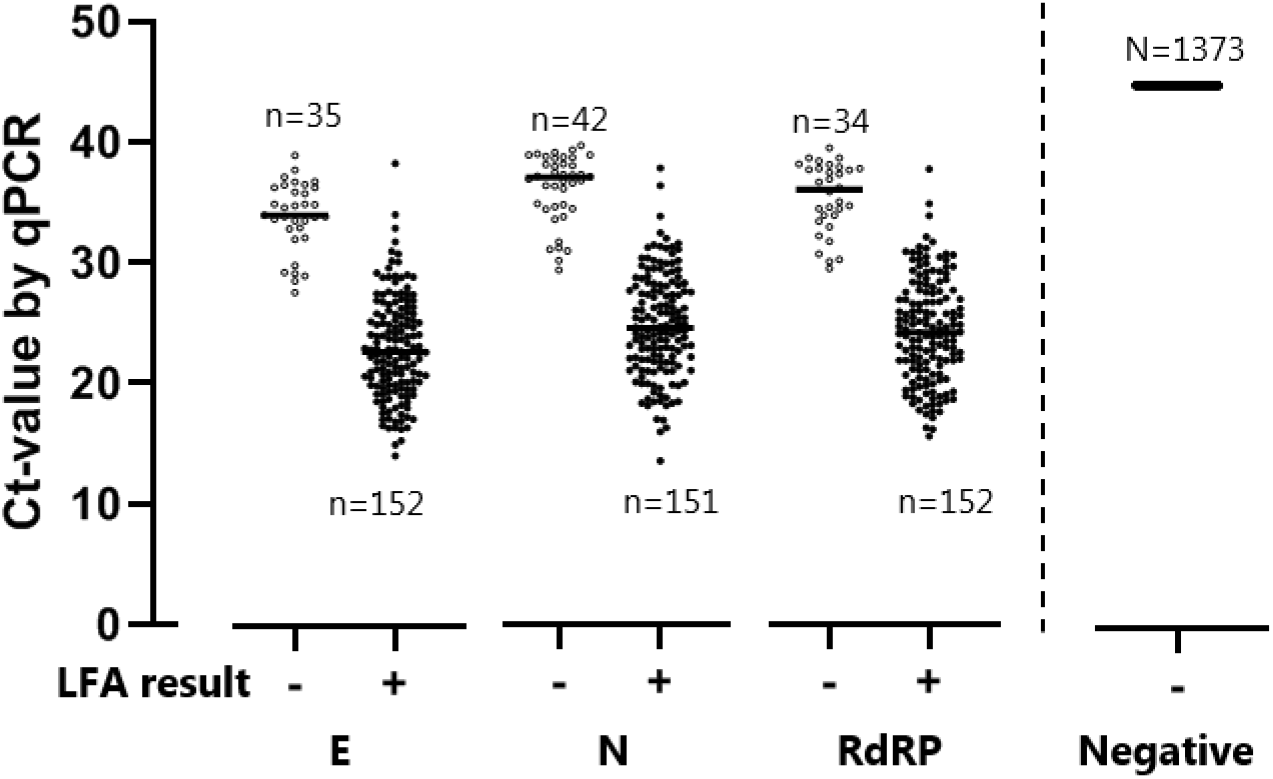
PCR and LFA results of all subjects. All PCR results for the three targets are shown by Ct value on the y-axis (left side: positive PCR results per target; right side: negative PCR results), grouped based on the LFA result on x-axis.

**Figure 2:**
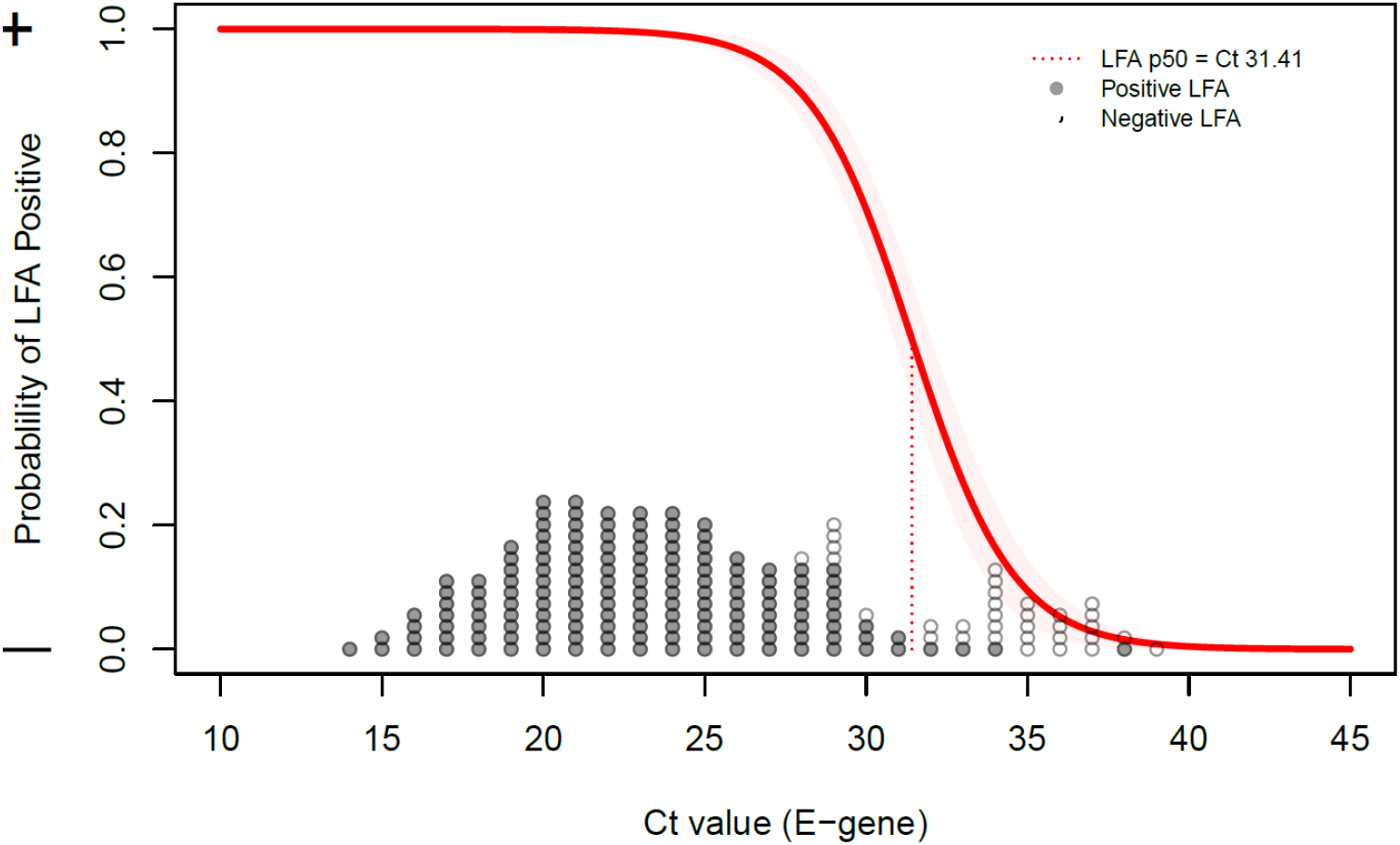
Association between Ct value and LFA test result. All dots reflect positive PCR results, shown on the x-axis at the observed Ct value of the E-gene. Grey dots reflect positive LFA samples, white dots negative LFA samples. The red line reflects the probability of a positive LFA based per Ct value, the red dotted line denotes the point where 50% of LFAs are expected to become positive.

### Relation with symptoms

The duration of symptoms was not associated with Ct-values (p = 0.46) or with the occurrence of false negative LFA results (p = 0.30) (Figure 3). When including only symptoms positively associated with COVID-19 (i.e., fever, chills, loss of taste/smell, muscle- or joint ache), duration of symptoms was weakly associated with Ct-values (p= 0.02), but no such association was found with LFA results (p= 0.45). Restricting analyses to individuals with symptoms for less than 7 days did not change sensitivity of LFA neither for all subjects positive with RT-qPCR (74.3%; 95%CI 64.6 – 82.4%) or when applying a cut-off at Ct>32 (97.4%; 95%CI 90.9 –99.7%).

**Figure 3.**
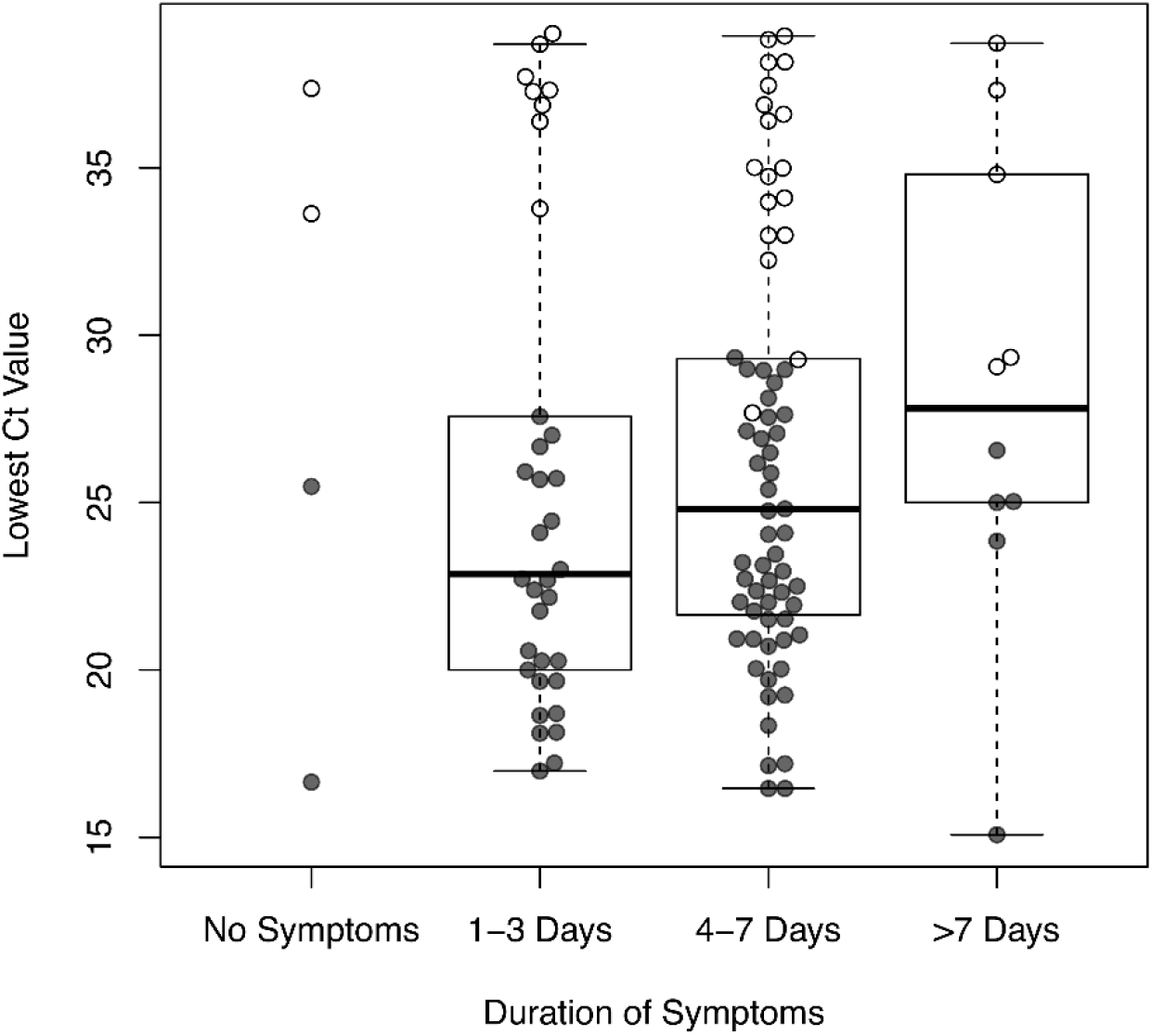
Ct value of positive subjects grouped by duration of symptoms. The dots represent individuals with positive PCR results shown on the y-axis based on the lowest observed Ct value in any of the three targets. The dots are groups based on the duration of symptoms. Grey dots represent positive LFA results, white dots negative LFA results.

## Discussion

In this real-life evaluation of the Panbio™ COVID-19 Ag rapid test in community-dwelling subjects with mild symptoms of respiratory tract infection, the assay reliably identified SARS-CoV-2 infected subjects with low Ct values by RT-qPCR (i.e. infections with a high viral load in nasopharyngeal samples). In our study cohorts specificity was 100%, overall sensitivity was 72.6% and 95.2% when using a Ct value of 32 as cut-off.

This new LFA test has not been evaluated extensively. The manufacturer reported a higher sensitivity (93.3%; 95CI 83.8-98.2), obtained in a high endemic setting in Brazil, testing individuals with symptoms for less than seven days only.^12^ In another cohort of 257 patients (both symptomatic and asymptomatic) enrolled at the emergency department and primary health care setting in Spain, overall sensitivity was 73.3%, and 86.5% among individuals with symptoms for less than seven days.^8^

In our study cohort, false negative results were observed only at high Ct values, i.e. with low viral load in nasopharyngeal material. This may occur very early in the infection (presymptomatic stage) before viral replication peaks, or in a late stage of infection when replication has decreased. Individuals with symptoms for more than 7 days may, therefore, be more likely to have a low viral load in nasopharyngeal swabs than those tested shortly after symptom onset. Indeed, a longer duration of symptoms associated with SARS-CoV-2 infection, was associated with higher Ct values, but no such association was observed between false negative LFA results and duration of symptoms. As a result, test sensitivity did not substantially increase when analyses were restricted to individuals with symptoms for less than a week. It must be noted that the study period corresponded with high prevalence of seasonal rhinovirus and other respiratory infections in the Netherlands,^13,14^ thereby possibly obscuring the association of upper respiratory tract symptoms with SARS-CoV-2 infection.

From a public healthcare perspective, missed infections with the LFA in patients with high Ct values in a late stage of infection may have limited impact, as these individuals are less likely to contribute to transmission. This is supported by findings that culturing of SARS-CoV-2 appeared not possible at Ct values above 29.^15^Yet, false negative LFA results were also observed among individuals with a very short duration of symptoms and high Ct values. A missed infection in the early stage may have consequences for transmission as these individuals may become infectious^16–18^. Furthermore, the majority of individuals in our cohort reported (mild) symptoms. Studies are needed to determine the sensitivity of this LFA test in asymptomatic subjects with detectable SARS-CoV-2 in RT-qPCR. These studies could also provide more insight in the performance of this LFA in the presymptomatic stage of infection.

For symptomatic persons requiring hospital admission a diagnostic test with high sensitivity is needed to establish a definitive clinical diagnosis.^19^ In this clinical context, we recommend to use RT-qPCR, as its sensitivity is superior to this LFA, allowing detection of individuals in the presymptomatic and late stage of infection which are both relevant in this context. However, in the context of community-based surveillance, tests need to identify symptomatic and asymptomatic infections in short time to stop onward spread. Transmission of SARS-CoV-2 is considered to occur mainly around symptom onset, when viral load peaks.^20,21^ This LFA, therefore, appears to reliably identify those patients that are most likely to contribute to onward transmission and could therefore be an essential new tool in our testing strategies. As the virus remains infectious in the assay sampling buffer (data not shown), biosafety should be taken into account when implementing the use of the Panbio Covid-19 LFA outside a laboratory setting. Considering the short turnaround time, user friendliness, opportunity for decentralized testing and low costs we believe that in the context of community-based surveillance of symptomatic individuals, these advantages outweighs the lower sensitivity of LFA compared to RT-qPCR. Modelling studies may provide further reassurance on the safety of such an approach.

## Data Availability

All available deidentified participant data can be made available upon request per email to the corresponding author.

## Author contributions

LMH, HG, BW designed the study. MB, AW, NR and RS provided counsel on study design and data interpretation. Sample collection and data analysis was performed by HG, BW and LMH at the UMC Utrecht and OR, JU, AR in LABHOH on Aruba. HG, BW and LMH drafted the manuscript. All authors contributed to critical revision of the manuscript.

## Acknowledgements

The authors would like to thank all participating employees from the GGD Utrecht and the department of Public Health in Aruba and the participating students from the UMCU for their help with data collection, as well as all participating volunteers.

## Data sharing statement

All deidentified participant available data can be made available upon request per email to the corresponding author.

